# Effectiveness of Kalevala Manual Therapy, Health Coaching, and Physiotherapy for Chronic Low Back Pain: A Randomized Controlled Trial (The INTEGRA Trial)

**DOI:** 10.64898/2026.10.13.26364897

**Authors:** Mika Venojärvi, Sanna Mattila-Rautiainen, Markku Partinen, Tiina Finsjö, Pekka Puska, Mikko Nenonen

## Abstract

**Background and aim:** Low back pain is the leading cause of years lived with disability (YLDs) globally. While physiotherapy (PT) is recommended as a first-line treatment, evidence regarding Kalevala Manual Therapy (KMT) remains limited. This study compared the effects of physiotherapy, KMT, and KMT combined with health coaching (KMT+HC) on pain, disability, and psychosocial outcomes in individuals with CLBP.

**Experimental procedure:** In this single-center, three-arm, parallel-group trial, 81 adults with non-specific CLBP (symptoms ≥ 12 weeks, STarT Back score > 3) were randomized (1:1:1) to receive PT (*n* = 27), KMT (*n* = 27), or KMT+HC (*n* = 27) over an 8-week intervention period. Primary outcomes were functional disability (Oswestry Disability Index, ODI) and pain intensity (Numeric Rating Scale, NRS). Secondary outcomes included work- and physical activity-related fear-avoidance beliefs (FABQ-W, FABQ-PA), depressive symptoms (BDI), quality of life (RAND-36), and patient-specific functional status (PSFS). Longitudinal changes were analyzed using linear mixed models.

**Results:** Statistically significant and generally clinically meaningful improvements over time were observed across the intervention groups for both ODI and NRS (both p < 0.001), with no significant between-group differences in the primary outcomes. No significant between-group differences were identified for the primary outcomes. A significant group-by-time interaction was observed for work-related fear-avoidance beliefs (FABQ-W; *p* = 0.005). Post hoc analyses revealed that KMT+HC achieved a significantly greater reduction in FABQ-W scores than PT (*p* = 0.002), with only the KMT+HC group exceeding the established minimal clinically important difference (MCID) threshold. Overall treatment satisfaction was high, with a mean score of at least 6.2 out of 7 and 100% protocol adherence were observed across all arms without any reported adverse events.

**Conclusion:** KMT and KMT combined with health coaching produced improvements in pain and functional disability comparable to guideline-based physiotherapy. Furthermore, the addition of structured health coaching to KMT significantly reduced work-related fear-avoidance beliefs, demonstrating the value of integrating behavioral support into manual therapy for comprehensive CLBP rehabilitation.

## 1. Introduction

Chronic low back pain (CLBP) is the leading cause of years lived with disability (YLDs) globally and represents a substantial global health burden and is associated with substantial healthcare utilization, reduced quality of life, work related disability, and economic burden [1]. Contemporary evidence indicates that CLBP should be understood within a biopsychosocial framework in which biological, psychological, and social factors interact to influence pain persistence and disability [2]. Consequently, modern treatment recommendations emphasize multimodal management strategies that combine physical, educational, and behavioral approaches rather than relying exclusively on passive treatments [3].

Physiotherapy remains a cornerstone of conservative CLBP management. Nevertheless, treatment responses vary considerably between individuals, and no single intervention has demonstrated universal superiority. In a multicenter randomized controlled trial (RCT), in individuals with CLBP, Cognitive Functional Therapy (CFT) produced significantly greater improvements in pain and disability than a group-based exercise and education program in individuals with CLBP [4]. In addition, Blanco-Giménez and co-workers demonstrated in a randomized controlled trial that combining manual therapy with exercise resulted in greater improvements in disability, kinesiophobia (fear of movement), self-efficacy, and pain-related outcomes than exercise alone in individuals with CLBP [5]. Adherence to prescribed home-based exercises may substantially influence the effectiveness of physiotherapy interventions. Previous research has shown that patients with declining adherence to home exercise recommendations showed poorer recovery and smaller improvements in pain and disability than patients who maintained or increased their exercise adherence during treatment. These findings suggest that treatment outcomes may be affected not only by the intervention itself but also by the patient’s commitment to the prescribed rehabilitation program [6].

Manual therapy comprises a variety of hands-on interventions, including joint mobilization, manipulation, and soft-tissue treatment techniques, and is widely used in the management of chronic musculoskeletal pain. Several randomized controlled trials and systematic reviews have reported beneficial effects on pain and functional outcomes, particularly when manual therapy is combined with exercise-based rehabilitation [7,8]. Kasimis et al. demonstrated that manual therapy combined with pain neuroscience education and motivational interviewing resulted in greater improvements in pain, disability, kinesiophobia, and functional performance than either manual therapy alone or home exercise in individuals with chronic non-specific low back pain [9]. Such findings support the view that manual interventions may be most effective when delivered within a broader biopsychosocial framework.

Health coaching has emerged as a patient-centered strategy that seeks to enhance self-management, adherence to rehabilitation, physical activity, and long-term behavioral change. Rather than focusing solely on symptom reduction, health coaching aims to strengthen self-efficacy and encourage sustainable lifestyle modifications. Previous studies have suggested that treatment adherence influences recovery in chronic low back pain [6], while interventions incorporating motivational and behavioural support may further enhance functional outcomes and self-management [9]. Consequently, combining health coaching with manual therapy may provide synergistic benefits by coupling symptom relief with strategies that promote long-term self-management.

Kalevala Manual Therapy (KMT), traditionally known as Kalevala bone setting, is a Finnish manual therapy method characterized by whole-body soft-tissue treatment and non-manipulative techniques. Despite its long history and widespread use in Finland, only few controlled studies have been done to evaluate its effectiveness. One of the earliest randomized studies was conducted by Hemmilä and coworkers, who compared traditional bone-setting treatment with physiotherapy in patients with chronic back pain and reported favorable long-term outcomes following bone-setting treatment [10]. Subsequently, Zaproudina et al. investigated physiological correlates of low back pain using plantar infrared thermography and demonstrated associations between thermal patterns and low back pain characteristics [11]. Later, Zaproudina et al. conducted a randomized pilot trial evaluating traditional bone-setting treatment in chronic low back pain and reported clinically meaningful improvements in pain and function, supporting the need for larger controlled trials [12].

The World Health Organization (WHO) recently published evidence-based guidelines for the non-surgical management of chronic primary low back pain [13]. These recommendations emphasize person-centered care and advocate interventions that support physical activity, patient education, self-management, exercise therapy, and psychological or behavioral approaches. The WHO guideline specifically highlights the importance of integrating biological, psychological, and social dimensions of care while discouraging excessive reliance on passive treatment modalities lacking evidence for long-term effectiveness [13]. These recommendations are consistent with the growing international consensus that sustainable improvements in CLBP are most likely achieved through active participation in rehabilitation and behavioral change [3,13].

Previous studies have reported similar beneficial effects of Kalevala Manual Therapy on pain and disability [10, 12]. Nevertheless, randomized controlled trials comparing KMT with current guideline-based physiotherapy interventions are still lacking. Furthermore, no previous randomized study has examined whether combining KMT with structured health coaching can enhance treatment outcomes through improved self-management, increased physical activity, and reductions in fear-avoidance behavior.

We hypothesized that all interventions would improve pain and disability and examined whether the addition of health coaching to Kalevala Manual Therapy was associated with different outcomes compared with Kalevala Manual Therapy alone and physiotherapy.

## 2. Materials and Methods

### 2.1 Study design and population

This study was a single-center, three-arm, parallel-group randomized controlled trial conducted at the University of Eastern Finland. The intervention period lasted 8 weeks. Participants were randomized in a 1:1:1 ratio to physiotherapy (PT), Kalevala Manual Therapy (KMT), or Kalevala Manual Therapy combined with health coaching (KMT+HC).

### 2.2 Participant eligibility criteria

Inclusion criteria were age ≥18 years, chronic low back pain lasting at least 12 weeks, STarT Back score >3 [14] and ability to participate in the intervention protocol. Exclusion criteria included pregnancy, pacemaker, metallic prosthesis, rheumatic disease, severe spinal deformity, previous spinal surgery, acute disc herniation and severe sciatica.

A total of 81 adults with chronic non-specific low back pain who met the eligibility criteria were recruited from the municipalities of Kuopio, Siilinjärvi, and Leppävirta (Figure 1).

**Figure 1.**
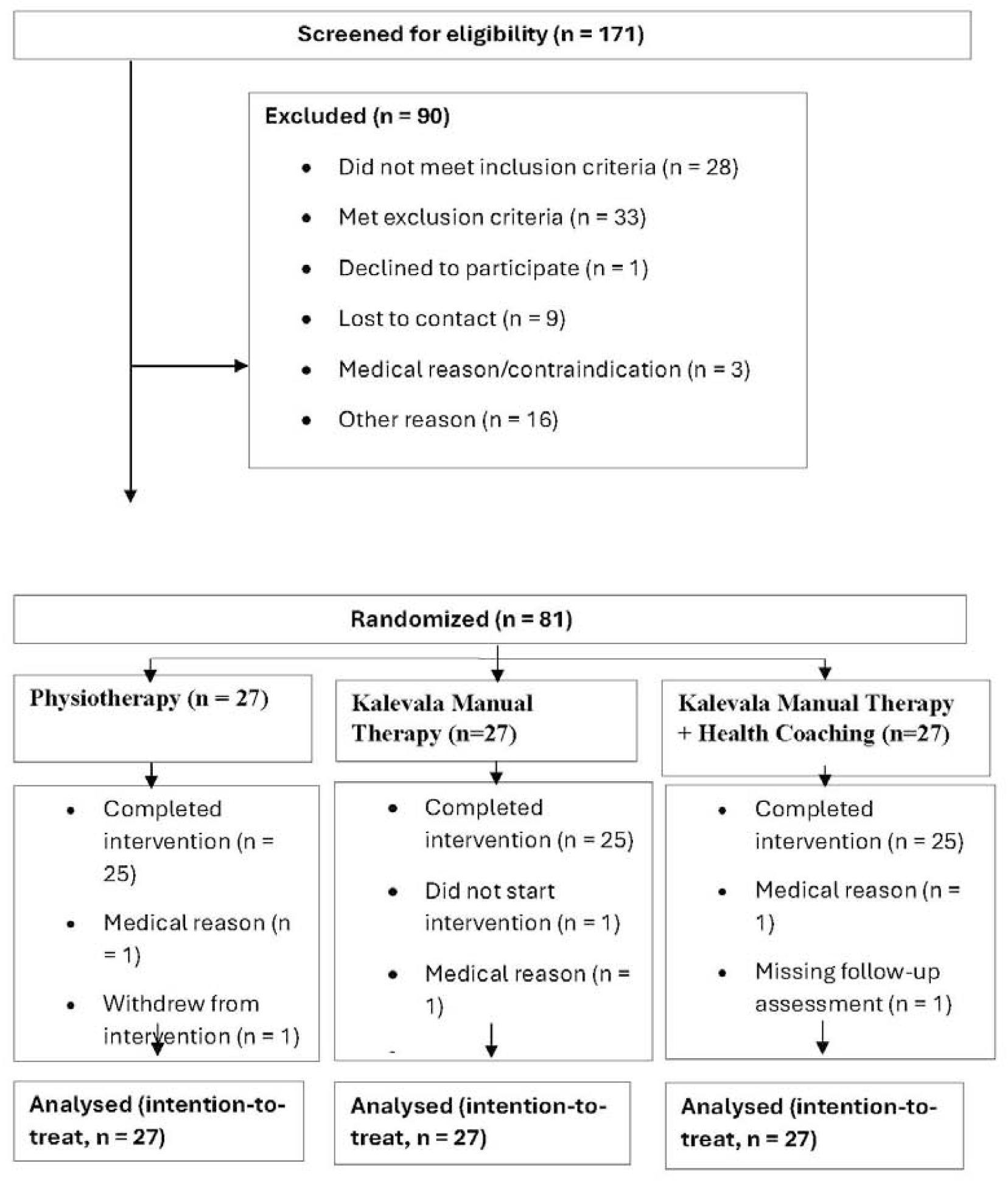
CONSORT flow diagram of participant recruitment, allocation, follow-up, and analysis. Participants were screened for eligibility, randomized to physiotherapy (PT), Kalevala Manual Therapy (KMT), or Kalevala Manual Therapy combined with Health Coaching (KMT+HC), and included in the intention-to-treat analyses.

### 2.3 Interventions

The physiotherapy intervention was delivered by a certified physiotherapist with more than 10 years of experience with pain patients. Kalevala Manual Therapy is a traditional Finnish manual therapy approach derived from indigenous bone-setting traditions and later formalized through structured education and standardized clinical practice. The treatments were delivered by three formally trained practitioners holding the protected professional title of “Kalevalainen jäsenkorjaaja”, each with more than 10 years of clinical experience. Health coaching was conducted by exercise medicine students under the supervision of the principal investigator and in accordance with the study protocol.

#### 2.3.1 Physiotherapy

The physiotherapy intervention consisted of three individual sessions delivered over 8 weeks (60 min at baseline and 30 min sessions at weeks three (3) and six (6), combined with a home exercise programme. The programme emphasized trunk stabilization, lumbopelvic control, hip strengthening, flexibility training, and progressive aerobic exercise. Participants were instructed to perform the exercises independently between visits and to gradually increase physical activity levels. A detailed description of the physiotherapy programme is provided in Supplementary Appendix 1.

#### 2.3.2 Kalevala Manual Therapy

Participants received three KMT sessions over eight weeks. Each session lasted approximately 90 minutes. The manual therapy intervention implemented in this study was a non-invasive tissue-based treatment designed to support natural mobility and functional movement. The treatments were administered by trained and certified practitioners of Kalevala manual therapy (*Kalevalainen jäsenkorjaus*). Prior to each session, an initial assessment was conducted to identify any contraindications and ensure the participant’s suitability for treatment.

The treatment progressed systematically from the lower body towards the upper body, covering the feet and ankles, lower legs, knees, thighs and hips, the sacral and spinal regions, upper extremities, and the head and cervical spine. Treatment techniques consisted of soft, slow, and tissue-oriented manual techniques aimed at supporting normal tissue mobility and fluid circulation. The techniques were applied in a controlled manner without sudden or forceful movements and did not include spinal or joint manipulation. Following treatment, participants received instructions regarding post-treatment self-monitoring and were advised to contact the research team if any unusual or persistent symptoms occurred.

#### 2.3.3 Kalevala Manual Therapy plus Health Coaching

Participants in the KMT+HC group received six structured telephone-based health coaching sessions (approximately 30 min each) during weeks 1, 2, 3, 5, 6, and 7 and three KMT sessions over eight weeks. The intervention was based on a biopsychosocial model of chronic low back pain and incorporated motivational interviewing and self-management support. Session content addressed pain education, goal setting, fear-avoidance beliefs, coping strategies, physical activity, ergonomic principles, behavioural change, and relapse prevention. Participants were also encouraged to increase daily physical activity and follow an individualized home-based exercise programme including walking, mobility, trunk stabilization, and strengthening exercises. A detailed description of the health coaching programme is provided in Supplementary Appendix 2.

### 2.4 Outcomes

Primary outcomes were disability measured with the modified Oswestry Disability Index (ODI) and pain intensity measured with the Numeric Rating Scale (NRS) [15,16]. Disability was assessed using the Oswestry Disability Index (ODI), a 10-item questionnaire measuring low back pain-related disability, with scores ranging from 0 to 100, where higher scores indicate greater disability [15]. Pain intensity was assessed using an 11-point Numeric Rating Scale (NRS; 0 = no pain, 10 = worst imaginable pain). Participants reported the lowest and highest pain intensity experienced during the preceding week. Secondary outcomes included the Beck Depression Inventory (BDI), Fear-Avoidance Beliefs Questionnaire (FABQ), modified short form of the Global Physical Activity Questionnaire (GPAQ), Patient-Specific Functional Scale (PSFS), modified Pittsburgh Sleep Quality Index (PSQI), and RAND 36-Item Health Survey (RAND-36). Detailed descriptions, scoring methods, and psychometric properties of these instruments have been reported previously and are referenced accordingly.

Depressive symptoms were assessed using the Beck Depression Inventory (BDI), a validated 21-item questionnaire commonly used in chronic pain and low back pain research to assess the severity of depressive symptoms [17,18]. Fear-avoidance beliefs were assessed using the Fear-Avoidance Beliefs Questionnaire (FABQ), which comprises 16 items and two subscales: physical activity (FABQ-Phys, 0-24) and work-related activities (FABQ-W, 0-42) [19,20]. Higher scores indicate greater fear-avoidance beliefs. The Minimal Clinically Important Difference (MCID) values are 4 points for FABQ-Phys and 7 points for FABQ-W. [20] Physical activity was assessed using a modified short-form GPAQ [21]. Participants reported the number of days per week engaging in light-intensity physical activity, moderate-intensity physical activity, as well as their average daily sitting time at weekdays. Functional ability was assessed using the Patient-Specific Functional Scale (PSFS), a patient-centered measure in which participants identify and rate activities that are difficult to perform because of their condition. Scores range from 0 to 10, with higher scores indicating better functional ability [22]. Sleep quality was assessed using the Pittsburgh Sleep

Quality Index (PSQI), a 19-item questionnaire evaluating subjective sleep quality and disturbances during the previous month. Higher scores indicate poorer sleep quality [23]. Health-related quality of life was assessed using the RAND 36-Item Health Survey (RAND-36), which measures perceived health across eight physical and mental health domains. Scores range from 0 to 100, with higher scores indicating better health-related quality of life [24].

#### 2.4.1 Participant feedback and treatment satisfaction

Following the intervention, participants completed a study-specific feedback questionnaire. Satisfaction with the intervention and perceived treatment effects were assessed using seven statements rated on a 7-point Likert scale (1 = strongly disagree, 7 = strongly agree). The statements evaluated whether the treatment period met participants’ expectations, perceived usefulness of the intervention, reduction in low back pain symptoms, improvement in functional ability, increased understanding of low back pain, overall satisfaction with the treatment, and willingness to recommend the intervention to others.

### 2.5 Sample size

A priori power analysis (α = 0.05, power = 0.80) indicated that 30 participants per group were required to detect a clinically meaningful between-group difference of 2 NRS points (SD = 3; Cohen’s d = 0.67). To account for potential attrition, the target sample size was set at 35 participants per group; however, 27 participants per group were ultimately enrolled.

### 2.6 Randomization

Randomization was performed using a concealed computer-generated allocation sequence with variable block sizes and stratification by sex and age (<44 years and ≥44 years). Group allocation was revealed after completion of baseline assessments. The trial was conducted according to CONSORT recommendations for non-pharmacological interventions.

### 2.7 Data collection

Outcome assessments were performed at baseline and immediately after the 8-week intervention. Adverse events, adherence and participant satisfaction were recorded throughout the study.

### 2.8 Statistical analyses

All statistical analyses were performed using IBM SPSS Statistics for Windows, version 29 (IBM Corp., Armonk, NY, USA). Continuous variables are presented as means and standard deviations (SDs) or estimated marginal means (EMMs) with 95% confidence intervals (CIs). Baseline differences between intervention groups were assessed using one-way analysis of variance (ANOVA) for normally distributed variables, the Kruskal-Wallis test for non-normally distributed variables, and chi-square tests for categorical variables. The primary outcomes were disability, assessed with the Oswestry Disability Index (ODI), and highest pain intensity, assessed using a Numeric Rating Scale (NRS). Secondary outcomes included Fear-Avoidance Beliefs Questionnaire physical activity (FABQ-PA) and work (FABQ-W) subscales, Beck Depression Inventory (BDI), Pittsburgh Sleep Quality Index (PSQI), sitting time and physical activity variables, Patient-Specific Functional Scale (PSFS) and RAND-36 health-related quality-of-life domains. Longitudinal changes in outcomes were analyzed using linear mixed models with Group, Time, and the Group × Time interaction specified as fixed effects. Estimated marginal means and 95% confidence intervals were derived from the fitted models. For outcomes demonstrating a significant Group × Time interaction, change scores (follow-up minus baseline) were calculated and compared between groups using one-way analysis of variance (ANOVA). Bonferroni-adjusted post hoc comparisons were performed to identify pairwise group differences. To examine factors associated with disability, an additional adjusted linear mixed model was fitted with ODI as the dependent variable and body weight, pain intensity (NRS), fear-avoidance beliefs related to work (FABQ-W), depressive symptoms (BDI), and sleep quality (PSQI) entered as covariates. Treatment satisfaction and participant feedback were assessed using a seven-item questionnaire. Internal consistency of the questionnaire was evaluated using Cronbach’s alpha, and between-group differences were analyzed using Kruskal-Wallis tests. Statistical significance was set at *p* < 0.05 for all analyses.

### 2.9 Ethical considerations

The study was conducted in accordance with the ethical principles of the Declaration of Helsinki. Ethical approval was obtained from the Regional Medical Research Ethics Committee of the Eastern Finland Collaborative Area (Pohjois-Savo Wellbeing Services County, Kuopio, Finland) on 27 February 2026 (reference no. 62/2026 [207/2025]). All participants received written and verbal information about the study and provided written informed consent before participation. The trial was prospectively registered in the ISRCTN registry (ISRCTN13717352).

## 3. Results

### 3.1 Participant flow and baseline characteristics

A total of 171 individuals were screened for eligibility, of whom 81 were randomized to physiotherapy (PT, n = 27), Kalevala Manual Therapy (KMT, n = 27), or Kalevala Manual Therapy combined with health coaching (KMT+HC, n = 27) (Figure 1). During the intervention period, one participant in the PT group withdrew, one participant in the KMT group did not start the intervention, and one participant in the KMT+HC group did not complete the follow-up assessment. Longitudinal analyses were performed using linear mixed models, which included all available observations from randomized participants. (Figure 1)

Baseline demographic and clinical characteristics are presented in Table 1. The three study groups were generally comparable at baseline. No statistically significant between-group differences were observed in age, height, ODI score, NRS score, STarT Back score, work category, medication category, or pain duration category (*p* > 0.05). Body weight was the only variable that differed significantly between the groups (*p* = 0.029). Mean body weight was greater in the KMT group (91.5 ± 19.8 kg) than in the PT group (81.1 ± 15.0 kg) and the KMT+HC group (80.8 ± 13.6 kg).

**Table 1.** Baseline characteristics of participants in physiotherapy (PT), Kalevala Manual Therapy (KMT), and Kalevala Manual Therapy combined with Health Coaching (KMT+HC) groups.

| Variable | PT (n=27) | KMT (n=27) | KMT+HC (n=27) | p-value |
| --- | --- | --- | --- | --- |
| Age, years | 58.2 ± 14.6 | 54.4 ± 13.6 | 54.3 ± 13.5 | 0.497 |
| Female n (%) | 15 (55.6) | 14 (51.9) | 16 (59.3) | 0.861 |
| Height, cm | 169.0 ± 7.9 | 172.0 ± 12.7 | 171.6 ± 10.3 | 0.520 |
| Weight, kg | 81.1 ± 15.0 | 91.5 ± 19.8 | 80.8 ± 13.6 | 0.029 |
| STarT Back score | 5.52 ± 1.34 | 5.37 ± 1.21 | 5.07 ± 1.14 | 0.432 |
| ODI score, % | 34.9 ± 13.8 | 33.5 ± 11.6 | 29.9 ± 12.9 | 0.269 |
| NRS, highest pain | 7.52 ± 1.67 | 7.11 ± 1.89 | 7.63 ± 1.64 | 0.608 |
| Work category, n (%) |  |  |  | 0.135 |
| Sedentary work | 11 (40.7) | 11 (40.7) | 18 (66.7) |  |
| Physical work | 3 (11.1) | 2 (7.4) | 1 (3.7) |  |
| Healthcare/service work | 3 (11.1) | 6 (22.2) | 0 (0.0) |  |
| Retired | 10 (37.0) | 8 (29.6) | 8 (29.6) |  |
| Medication category, n (%) |  |  |  | 0.309 |
| NSAIDs | 13 (48.1) | 6 (22.2) | 8 (29.6) |  |
| Opioids | 2 (7.4) | 3 (11.1) | 2 (7.4) |  |
| Other medication | 11 (40.7) | 14 (51.9) | 11 (40.7) |  |
| No medication | 1 (3.7) | 4 (14.8) | 6 (22.2) |  |
| Pain duration, n (%) |  |  |  | 0.360 |
| < 1 year | 6 (22.2) | 3 (11.1) | 4 (14.8) |  |
| 1–5 years | 13 (48.1) | 9 (33.3) | 13 (48.1) |  |
| > 5 years | 8 (29.6) | 15 (55.6) | 10 (37.0) |  |

Data are presented as mean ± standard deviation (SD) or frequency (%). Age, height, and weight were compared using one-way analysis of variance (ANOVA). ODI, Oswestry Disability Index; NRS, Numeric Rating Scale, NSAID, non-steroidal anti-inflammatory drug and STarT Back scores were compared using the Kruskal-Wallis test due to deviations from normality. Categorical variables (sex, work category, medication category, and pain duration) were compared using Pearson's chi-square test. Statistical significance was set at $p < 0.05$ . ODI baseline data were available for 80 participants (PT n=26, KMT n=27, KMT+HC n=27)

### 3.2 Primary outcomes Disability

ODI scores decreased significantly over the 8-week intervention period, from a mean of 32.8 at baseline to 21.4 at follow-up (p<0.001). The reduction was observed in all three intervention groups. However, neither the main effect of treatment group nor the group-by-time interaction was statistically significant, indicating no evidence of differential improvement in disability between PT, KMT, and KMT + HC (Figure 2). The magnitude of improvement reached the proposed minimal clinically important difference (MCID) in the PT and KMT+HC groups and was marginally below this threshold in the KMT group.

**Figure 2.**
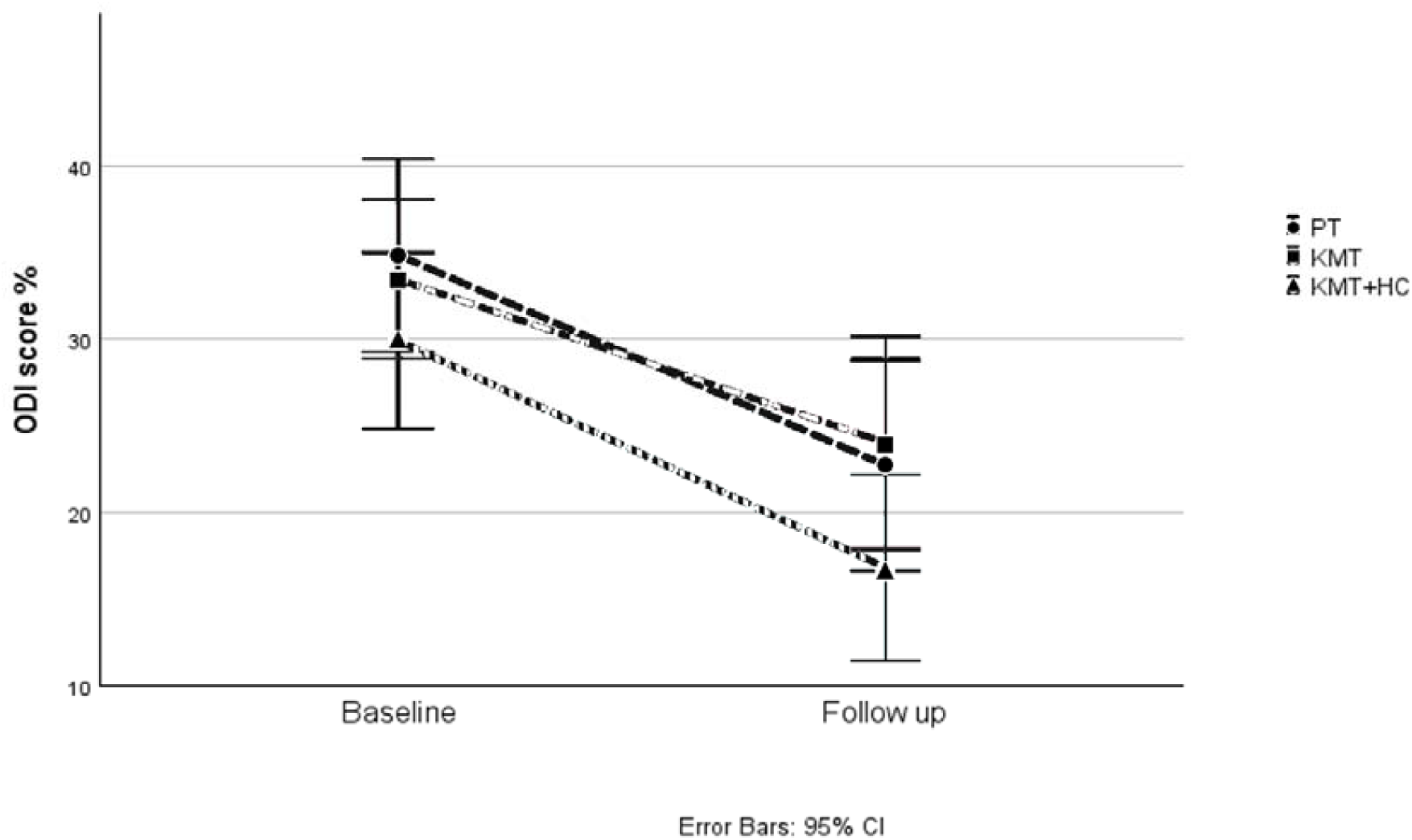
Estimated marginal means (EMMs) for highest pain scores (NRS) at baseline and follow-up in the physiotherapy (PT), Kalevala Manual Therapy (KMT), and Kalevala Manual Therapy combined with Health Coaching (KMT+HC) groups. Values are presented as means with 95% confidence intervals

#### Pain intensity

Highest weekly pain intensity, measured using the NRS, decreased significantly from 7.42 at baseline to 5.27 at follow-up (p<0.001). Pain reductions were observed in all three groups. No statistically significant between-group differences or group-by-time interaction effects were detected (Figure 3). The reduction in NRS scores reached the proposed MCID in the KMT and KMT + HC groups, whereas the improvement in the PT group was slightly below this threshold.

**Figure 3.**
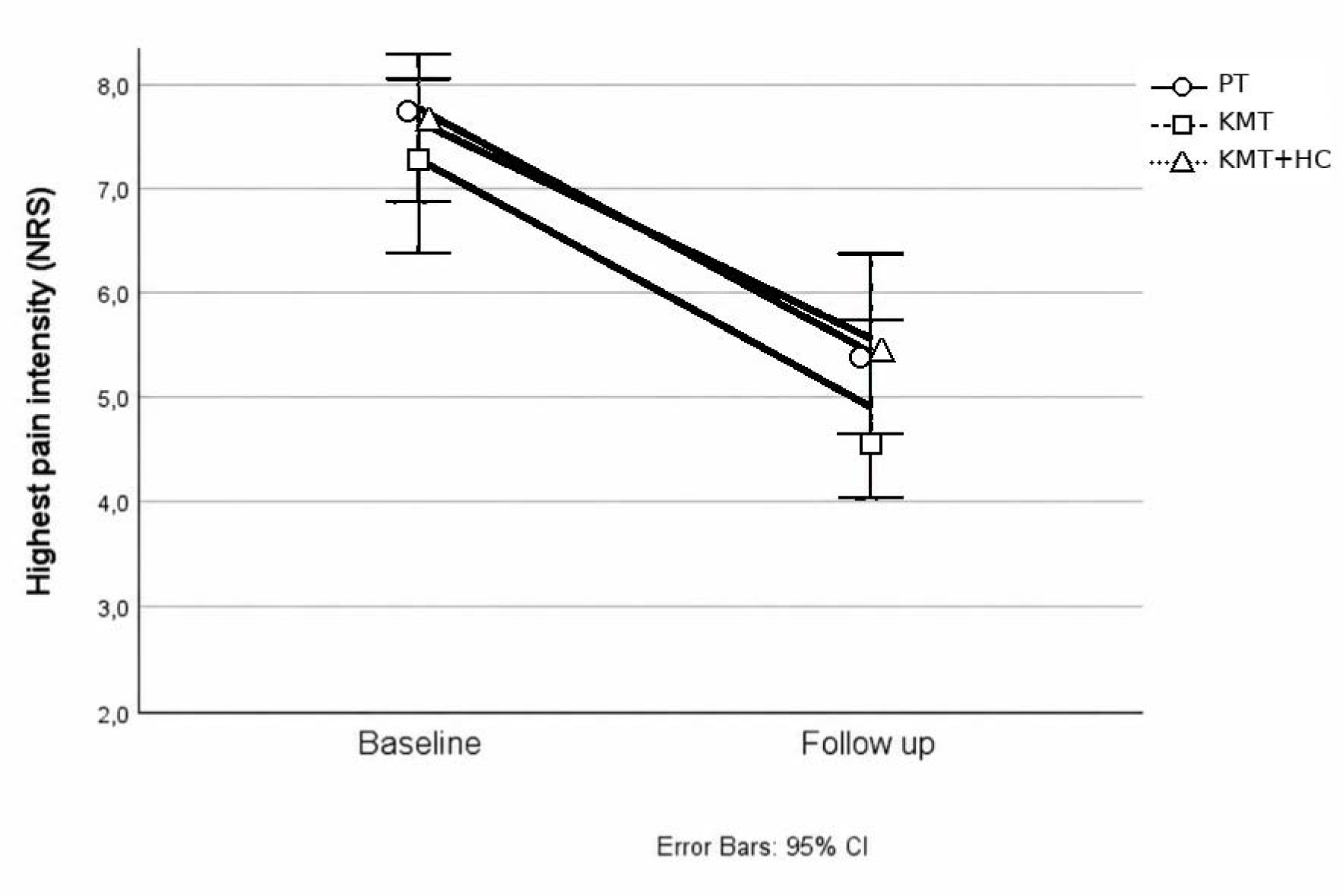
Estimated marginal means (EMMs) for ODI scores at baseline and follow-up in physiotherapy (PT), Kalevala Manual Therapy (KMT), and Kalevala Manual Therapy combined with Health Coaching (KMT+HC) groups. Values are presented as means with 95% confidence intervals.

### 3.3 Factors associated with disability

In the adjusted linear mixed model, higher pain intensity (NRS highest pain), greater fear-avoidance beliefs related to work (FABQ-W), and more severe depressive symptoms (BDI) were independently associated with higher ODI scores. All associations were statistically significant (p<0.05). Body weight and sleep quality (measured by PSQI) were not significant predictors of disability. After adjustment for these covariates, ODI scores continued to decrease over time (p=0.039). Neither the treatment group effect nor the group-by-time interaction was statistically significant, further indicating comparable changes in disability across the three intervention groups (Table 2).

**Table 2.** Adjusted linear mixed model analysis of ODI score.

| Variable | F | p-value |
| --- | --- | --- |
| Treatment group | 1.35 | 0.266 |
| Time | 4.36 | 0.039 |
| Treatment group × Time | 0.58 | 0.564 |
| Body weight | 0.90 | 0.345 |
| NRS highest pain | 25.81 | <0.001 |
| FABQ-W | 5.60 | 0.020 |
| BDI | 4.13 | 0.045 |
| PSQI | 0.78 | 0.380 |

Adjusted linear mixed model including body weight, NRS (Numeric Rating Scale) highest pain, FABQ-W (Fear-Avoidance Beliefs Questionnaire Work), BDI (Beck Depression Inventory ) and PSQI ( Pittsburgh Sleep Quality Index ) as covariates. Statistical significance was set at $p < 0.05$ .

### 3.4 Secondary outcomes

Significant improvements over time were observed in multiple secondary outcomes, including fear-avoidance beliefs, depressive symptoms, pain-related measures, social functioning, sitting time, and Patient-Specific Functional Scale. However, most secondary outcomes did not demonstrate statistically significant differences between groups. A significant group-by-time interaction was observed for FABQ-W (*p* = 0.005) (Table 3). Post hoc comparisons showed that participants in the KMT+HC group experienced a significantly greater reduction in FABQ-W scores than those in the PT group (*p* = 0.002). No statistically significant differences were observed between the PT and KMT groups or between the KMT and KMT+HC groups. The greatest improvement in FABQ-W was observed in the KMT+HC group (−8.0 points), exceeding the proposed MCID of 7 points. In contrast, improvements in the KMT (−4.2 points) and PT (−0.7 points) groups did not reach the MCID threshold.

**Table 3.** Estimated marginal means of secondary outcomes at baseline and follow-up.

| **Outcome** | **PT B** | **PT F** | **KMT B** | **KMT F** | **KMT+HC B** | **KMT+HC F** |
| --- | --- | --- | --- | --- | --- | --- |
| FABQ-PA | 7.1 (5.1-9.1) | 4.4 (2.3-6.4) | 8.2 (6.2-10.2) | 5.5 (3.5-7.6) | 7.2 (5.2-9.2) | 4.2 (2.1-6.2) |
| FABQ-W | 19.1 (15.0-23.3) | 18.2 (13.9-22.4) | 21.6 (17.5-25.8) | 17.4 (13.2-21.7) | 17.9 (13.7-22.1) | 9.9 (5.7-14.1) |
| BDI | 7.7 (5.7-9.7) | 6.8 (4.8-8.9) | 8.4 (6.3-10.4) | 5.9 (3.8-7.9) | 7.7 (5.6-9.7) | 5.2 (3.1-7.3) |
| PSQI | 8.9 (7.5-10.4) | 9.4 (7.9-10.9) | 7.8 (6.3-9.3) | 8.6 (7.1-10.1) | 8.4 (6.9-9.8) | 7.6 (6.1-9.1) |
| RAND-36 Physical Functioning | 64.6 (56.3-72.9) | 70.6 (62.3-78.9) | 67.3 (59.0-75.6) | 68.1 (59.8-76.4) | 70.2 (61.9-78.5) | 75.2 (66.9-83.5) |
| RAND-36 Role Physical | 35.6 (21.1-50.1) | 52.9 (38.4-67.4) | 32.7 (18.2-47.2) | 49.0 (34.5-63.5) | 57.7 (43.2-72.2) | 66.3 (51.9-80.8) |
| RAND-36 Role Emotional | 64.1 (49.7-78.5) | 72.9 (58.2-87.6) | 68.0 (53.5-82.4) | 75.5 (60.9-90.2) | 73.1 (58.6-87.5) | 83.2 (68.5-97.8) |
| RAND-36 Emotional Well-being | 72.9 (65.6-80.2) | 71.7 (64.4-79.0) | 74.1 (66.8-81.4) | 75.7 (68.4-83.0) | 73.8 (66.5-81.2) | 74.0 (66.7-81.3) |
| RAND-36 Social Functioning | 70.7 (62.0-79.3) | 77.9 (69.2-86.6) | 72.1 (63.4-80.8) | 77.9 (69.2-86.6) | 76.9 (68.2-85.6) | 89.1 (80.3-97.8) |
| RAND-36 Pain | 45.5 (38.2-52.8) | 54.3 (47.1-61.6) | 42.7 (35.4-50.0) | 55.7 (48.4-62.9) | 51.5 (44.3-58.8) | 62.5 (55.1-69.9) |
| RAND-36 General Health | 58.9 (51.6-66.1) | 56.0 (48.7-63.2) | 51.0 (43.7-58.2) | 54.6 (47.4-61.9) | 62.1 (54.9-69.3) | 65.6 (58.3-72.9) |
| Sitting time | 313.0 (248.2-377.7) | 269.4 (203.5-335.3) | 338.0 (273.2-402.7) | 291.9 (226.0-357.8) | 410.4 (345.6-475.1) | 335.6 (269.7-401.5) |
| Light physical activity | 419.3 (244.0-594.5) | 451.6 (270.1-633.1) | 397.8 (222.5-573.0) | 305.1 (120.2-490.0) | 364.7 (189.4-540.0) | 422.6 (241.1-604.1) |
| Moderate physical activity | 171.8 (83.5-260.2) | 173.7 (82.0-265.4) | 106.8 (18.5-195.2) | 156.3 (62.7-249.9) | 121.3 (33.0-209.6) | 84.7 (-7.0-176.4) |
| PSFS | 14.7 (12.5-17.0) | 20.1 (17.7-22.4) | 14.6 (12.4-16.9) | 18.1 (15.8-20.5) | 14.4 (12.2-16.7) | 21.8 (19.5-24.2) |

B. Linear mixed model p-values
| Outcome | Time | Group | Time×Group |
| --- | --- | --- | --- |
| FABQ-PA | <0.001 | 0.607 | 0.944 |
| FABQ-W | <0.001 | 0.103 | 0.005 |
| BDI | <0.001 | 0.808 | 0.204 |
| PSQI | 0.729 | 0.395 | 0.240 |
| RAND-36 Physical Functioning | 0.117 | 0.526 | 0.660 |
| RAND-36 Role Physical | 0.005 | 0.033 | 0.728 |
| RAND-36 Role Emotional | 0.050 | 0.546 | 0.974 |
| RAND-36 Emotional Well-being | 0.936 | 0.848 | 0.859 |
| RAND-36 Social Functioning | <0.001 | 0.229 | 0.478 |
| RAND-36 Pain | <0.001 | 0.100 | 0.828 |
| RAND-36 General Health | 0.437 | 0.065 | 0.252 |
| STarT Back | <0.001 | 0.250 | 0.928 |
| Sitting time | <0.001 | 0.156 | 0.615 |
| Light physical activity | 0.989 | 0.718 | 0.578 |
| Moderate physical activity | 0.890 | 0.345 | 0.614 |
| PSFS total | <0.001 | 0.361 | 0.218 |

Values are presented as estimated marginal means with 95% confidence intervals. P-values are reported for time, group, and time × group effects from linear mixed models. A. Estimated marginal means (EMM) with 95% confidence intervals. Statistical significance was set at $p < 0.05$ . PT, physiotherapy; KMT, Kalevala Manual Therapy; KMT+HC, Kalevala Manual Therapy combined with Health Coaching; B, baseline; F, follow-up; EMM, estimated marginal means; CI, confidence interval; FABQ-PA, Fear-Avoidance Beliefs Questionnaire—Physical Activity; FABQ-W, Fear-Avoidance Beliefs Questionnaire—Work; BDI, Beck Depression Inventory; PSQI, Pittsburgh Sleep Quality Index; RAND-36, 36-item RAND Health Survey; STarT Back, STarT Back Screening Tool; PSFS, Patient-Specific Functional Scale

### 3.5 Treatment satisfaction, adherence, and additional treatment

Of the 81 participants, 71 provided complete feedback questionnaire data and were included in the reliability analysis. The seven-item feedback questionnaire demonstrated good internal consistency, with a Cronbach’s alpha of 0.868. Overall participant evaluations were positive. The highest mean ratings were observed for recommending the intervention to others (6.46 ± 0.71) and overall satisfaction with treatment (6.20 ± 0.98). Participants also reported that the treatment was beneficial (6.15 ± 0.98), met their expectations (5.61 ± 1.22), increased their understanding of low back pain (5.51 ± 1.41), reduced symptoms (5.48 ± 1.42), and improved functional capacity (5.38 ± 1.35).

Among participants who completed the intervention, adherence to the planned treatment sessions was 100% in all groups. Minor scheduling deviations from the planned treatment schedule were observed in four participants (16%) in the PT group and one participant (4%) in the KMT group, reflecting treatment sessions that were conducted approximately one week earlier or later than originally scheduled. No treatment schedule deviations were observed in the KMT+HC group.

Of the 24 participants who reported home exercise adherence in the PT group, 45.8% performed the prescribed exercises 4-5 times per week and 20.8% exercised daily. In addition, 25.0% reported exercising 2-3 times per week, while no participant reported exercising less than once per week, once per week, or not at all.

No significant differences were observed between the PT, KMT, and KMT+HC groups in any feedback questionnaire item according to Kruskal-Wallis tests (all p > 0.05). Overall satisfaction, perceived treatment benefit, symptom relief, functional improvement, and willingness to recommend the intervention were comparable across groups (Table 4).

**Table 4.** Participant feedback and treatment satisfaction after the intervention.

| Outcome (1-7 Likert scale) | PT Mean ± SD | KMT Mean ± SD | KMT+HC Mean ± SD | p-value |
| --- | --- | --- | --- | --- |
| Intervention met expectations | 5.4 ± 1.4 | 5.7 ± 1.2 | 5.7 ± 1.1 | >0.05 |
| Low back pain symptoms were reduced | 5.4 ± 1.67 | 5.9 ± 1.00 | 5.2 ± 1.5 | >0.05 |
| Intervention was beneficial | 6.0 ± 1.3 | 6.4 ± 0.7 | 6.1 ± 0.9 | >0.05 |
| Functional ability improved | 5.2 ± 1.6 | 5.7 ± 1.0 | 5.2 ± 1.4 | >0.05 |
| Understanding of low back pain increased | 5.3 ± 1.7 | 5.7 ± 1.4 | 5.6 ± 1.2 | >0.05 |
| Overall satisfaction with treatment | 6.0 ± 1.3 | 6.3 ± 0.8 | 6.4 ± 0.8 | >0.05 |
| Would recommend intervention to others | 6.5 ± 0.7 | 6.7 ± 0.5 | 6.2 ± 0.8 | >0.05 |

Values are mean ± SD. Group differences were assessed using Kruskal-Wallis tests. No statistically significant differences were observed between groups (all $p > 0.05$ ). PT = physiotherapy; KMT = Kalevala Manual Therapy; KMT+HC = Kalevala Manual Therapy plus health coaching.

After excluding missing responses, most participants reported not using any additional treatment during the study period. In PT, 4 participants (16.0%) reported receiving manual therapy, 20 (80.0%) reported no additional treatment, and 1 (4.0%) reported other treatment. In KMT, 4 participants (17.4%) reported manual therapy, 17 (73.9%) no additional treatment, and 2 (8.7%) other treatment. In KMT+HC, no participants reported manual therapy, while 23 (92.0%) reported no additional treatment and 2 (8.0%) reported other treatment.

Adverse events were monitored throughout the intervention. No treatment-related adverse events or unintended effects were reported by participants in any of the study groups, including PT, KMT, and KMT+HC.

## 4. Discussion

The present randomized controlled trial demonstrated significant improvements in disability and pain intensity in all three intervention groups during the 8-week follow-up period. Participants receiving physiotherapy, Kalevala Manual Therapy, and Kalevala Manual Therapy combined with health coaching showed improvements in both ODI and NRS scores over time. Although clinically meaningful improvements were generally observed, the magnitude of change varied across outcomes and intervention groups. Improvements in ODI reached the proposed MCID in the PT and KMT+HC groups, while the improvement in the KMT group was only marginally below the threshold. Similarly, reductions in NRS reached the MCID in the KMT and KMT+HC groups, whereas the improvement in the PT group fell slightly short of the recommended threshold. Despite these differences, no significant between-group differences were observed for the primary outcomes, indicating that the three interventions produced broadly comparable effects on pain and disability. These findings indicate that physiotherapy, KMT, and KMT combined with health coaching can achieve clinically meaningful improvements in pain and disability among individuals with chronic low back pain.

These findings are consistent with previous randomized controlled trials demonstrating clinically meaningful improvements in pain and disability following conservative management of chronic low back pain. Kasimis et al. reported superior improvements when manual therapy was combined with pain neuroscience education and motivational interviewing compared with manual therapy alone, supporting the potential added value of behavioural and educational components alongside manual treatment [9]. Similarly, Zafereo et al. found significant improvements in disability and pain intensity over a 12-week period in both physiotherapy-based intervention groups, although no significant between-group differences were observed, which closely resembles the pattern identified in the present study [25]. Furthermore, a randomized pilot study by Zaproudina et al. demonstrated that traditional bone setting, a manual therapy approach related to Finnish folk medicine traditions, was associated with improvements in chronic low back pain and disability, further supporting the potential effectiveness of manual treatment approaches in this patient population [12]. Collectively, these findings suggest that manual therapy, physiotherapy, and multimodal interventions incorporating behavioural support may all contribute to meaningful clinical improvements in individuals with chronic low back pain.

Psychosocial factors and disability

The adjusted analyses supported a biopsychosocial interpretation of disability. Higher pain intensity, stronger fear-avoidance beliefs related to work, and more pronounced depressive symptoms were independently associated with higher ODI scores, whereas body weight and PSQI score were not significant predictors. The non-significant associations for body weight and PSQI should be interpreted cautiously, particularly given the relatively small sample size and the short follow-up period. They do not exclude the possibility that these factors may influence disability in larger or longer-term studies.

Similar findings have been reported in populations with chronic non-specific low back pain, where fear-avoidance beliefs have been identified as important predictors of disability and long-term outcomes [26,27]. Furthermore, psychosocial factors such as catastrophizing, kinesiophobia, and depressive symptoms have been reported to contribute substantially to disability beyond the direct effects of pain intensity [27,28].

Regarding secondary outcomes, the most notable finding was the greater reduction in work-related fear-avoidance beliefs among participants receiving KMT combined with health coaching compared with those receiving physiotherapy alone. FABQ-W was the only secondary outcome demonstrating both a significant group-by-time interaction and an improvement exceeding the proposed MCID [9,26]. Given the established role of fear-avoidance beliefs in disability and long-term outcomes in chronic low back pain, this finding may suggest that the combination of manual treatment and behavioural support provides added benefits in addressing maladaptive pain-related beliefs [9,26].

Improvements were also observed in depressive symptoms, selected domains of health-related quality of life, and self-reported ability to perform personally meaningful activities, as reflected by PSFS scores [22]. These findings are consistent with previous studies demonstrating that physiotherapy, manual therapy, and multimodal interventions can positively influence not only pain and disability but also broader psychosocial and functional outcomes in individuals with chronic low back pain [5,9].

In contrast, no significant changes were observed in sleep quality, emotional well-being, general health perceptions, physical functioning, or physical activity levels. While some previous multimodal interventions have reported broader psychosocial benefits [5,9], the absence of change in these outcomes in the present study may be related to the relatively short intervention period, differences in intervention content, or limited potential for improvement due to favourable baseline levels.

### Treatment satisfaction and adherence

Treatment satisfaction was high in all three groups, and participants generally perceived the interventions as beneficial and acceptable. Adherence to the scheduled treatment sessions was excellent, and no adverse events were reported. These findings support the feasibility and acceptability of KMT in this study population.

Nevertheless, treatment satisfaction and adherence should be interpreted in the context of the trial design. Participants were not blinded to treatment allocation, all interventions involved regular contact with study personnel, and the sample was recruited from a single geographical area. These factors may have contributed to positive treatment perceptions. In addition, some participants reported receiving additional treatment during the intervention period, which may have influenced the outcomes.

Treatment satisfaction was high across all intervention groups. Participants generally reported that the intervention met their expectations, was beneficial, improved their understanding of low back pain, and would be recommended to others. The questionnaire demonstrated good internal consistency, suggesting that these ratings reflected a coherent perception of treatment benefit. High satisfaction may be clinically relevant because patient acceptability is associated with treatment adherence, engagement in rehabilitation, and long-term self-management behaviors. Participant satisfaction was similarly high across the interventions, further supporting the acceptability and feasibility of KMT as a treatment option for chronic low back pain.

Importantly, study adherence was excellent across all groups. All participants who completed the study received the planned treatment sessions, and only minor treatment-scheduling deviations were observed. In addition, adherence to the home exercise programme was high in the PT group, with most participants reporting that they performed the prescribed exercises at least four times per week. This finding is noteworthy because previous research has shown that poorer adherence to home-based exercise is associated with less favorable recovery and smaller improvements in pain and disability in individuals with low back pain [6]. Together, these findings suggest that participant engagement and intervention fidelity were high regardless of treatment group, supporting the internal validity of the study and indicating that the observed outcomes are unlikely to be explained by poor treatment adherence.

The 100% attendance rate for scheduled treatment sessions should be interpreted in the context of participants who completed the intervention rather than the entire randomized sample. The high attendance may partly reflect the relatively limited number of scheduled treatment sessions, motivated trial participants, and regular contact with study personnel. Nevertheless, the high level of attendance indicates that the intervention protocols were feasible for participants who remained in the study.

Treatment outcomes in manual therapy may reflect not only the specific therapeutic techniques but also patient-related factors, practitioner expertise, expectations, therapeutic interaction, and the broader context in which care is delivered. These contextual factors may be particularly relevant to KMT, which is delivered as an individualized whole-body treatment involving substantial practitioner–patient interaction. The present study was not designed to separate the specific effects of manual techniques from contextual or practitioner-related effects. Future studies should therefore characterize and, where possible, evaluate treatment-context variables alongside clinical outcomes.

Future research should focus on both mechanisms and long-term outcomes. Follow-up analyses from the present trial, including six-month outcome data, will help determine whether the observed improvements are sustained over time. In addition, ongoing studies investigating immediate physiological and clinical responses to KMT may provide a better understanding of underlying treatment mechanisms. Such studies may clarify whether short-term changes in pain, mobility, autonomic nervous system activity, or other physiological parameters contribute to longer-term improvements in disability and quality of life. Larger multicentre trials with longer follow-up periods are warranted to confirm the present findings and establish the long-term effectiveness of KMT.

### Strengths and limitations

The strengths of this study include its randomized controlled design, the direct comparison of three active treatment approaches, comprehensive biopsychosocial outcome assessment, adjusted statistical analyses, and inclusion of patient satisfaction measures.

Several limitations should also be acknowledged. First, the sample size was smaller than planned and may have limited the ability to detect modest between-group differences. Second, the study was conducted at a single centre, which may limit generalizability. Third, participants and treatment providers could not be blinded, creating a possibility of expectancy and performance effects.

Fourth, the follow-up period was limited to eight weeks, so the durability of the improvements observed remains uncertain. Fifth, the study was not designed to test treatment equivalence or non-inferiority. Finally, the presence of additional treatments in some participants may have introduced contamination between groups.

### Implications and future research

The findings suggest that KMT may be a feasible and acceptable treatment option for individuals with chronic non-specific low back pain, producing short-term improvements similar to those observed following physiotherapy in the present study. The greater reduction in work-related fear-avoidance beliefs observed in the KMT+HC group further suggests that combining manual treatment with behavioural support may provide additional benefits for addressing psychosocial factors associated with persistent low back pain.

From a health-systems perspective, these findings are relevant to ongoing efforts to strengthen the evidence base for traditional, complementary, and integrative medicine. The WHO Global Traditional Medicine Strategy 2025-2034 emphasizes the importance of generating high-quality evidence, ensuring safety and quality, and supporting evidence-informed integration of traditional and complementary practices into health systems [29]. Within this context, the present study provides preliminary controlled evidence concerning KMT as a Finnish traditional manual therapy approach. However, the findings should not be interpreted as evidence of equivalence to physiotherapy or as sufficient justification for routine implementation.

Future studies should include larger multicentre samples, longer follow-up periods, and prespecified non-inferiority or equivalence designs. Further research is also needed to clarify the mechanisms underlying the observed effects and to determine whether improvements in pain, disability, and fear-avoidance beliefs are sustained over the longer term.

## Conclusion

In conclusion, physiotherapy, Kalevala Manual Therapy (KMT), and KMT combined with health coaching produced comparable improvements in pain and disability among individuals with chronic low back pain. No statistically significant between-group differences were observed for the primary outcomes. KMT combined with health coaching produced a greater reduction in work-related fear-avoidance beliefs than physiotherapy, and this reduction exceeded the proposed minimal clinically important difference. Larger studies with longer follow-up and formal equivalence or non-inferiority designs are needed to confirm these findings.

## Artificial Intelligence Use Statement

Microsoft Copilot was used to assist with language editing, manuscript organization, and preparation of tables and supplementary materials. All study design, data collection, statistical analyses, interpretation of the results, and final manuscript approval were performed by the authors. The authors take full responsibility for the content of this manuscript.

## Supplementary Material

Supplementary Appendix 1 contains the physiotherapy protocol and Supplementary Appendix 2 contains the health coaching protocol

## Author Contributions

Mika Venojärvi: Conceptualization, methodology, investigation, formal analysis, writing - original draft.

Mikko Nenonen: Conceptualization, review and editing.

Sanna Mattila-Rautiainen, Markku Partinen, Tiina Finsjö, and Pekka Puska: writing - review and editing

All authors read and approved of the final manuscript.

## Data Availability

All data produced in the present study are available upon reasonable request to the authors

## Acknowledgements

The authors would like to thank all study participants for their valuable contribution to this research. Special thanks are extended to M.Sc. (Health Sciences) Jenni Mikkanen for her exceptional work and commitment throughout the study. The authors also gratefully acknowledge Eveliina Rontu, Ida Grönvist, Kerttu Jääskeläinen, Emilia Komulainen, Pinja Filander, and Aada Tyrväinen, students of Sports and Exercise Medicine at the University of Eastern Finland, whose dedicated support in participant recruitment, study implementation, and data collection contributed substantially to the successful completion of the project.

## Trial Registration

This trial was prospectively registered in the ISRCTN registry (ISRCTN13717352) prior to participant recruitment.

## Appendix 1. Eight-week physiotherapy exercise programme

The programme comprised three supervised physiotherapy sessions (weeks 1, 3 and 6) combined with home exercise. Exercises progressed from trunk activation and mobility to strengthening and functional movement control. Aerobic exercise was included throughout the 8-week programme.

| Period | Main focus | Exercises and dosage | Aerobic exercise |
| --- | --- | --- | --- |
| Weeks 1–2 | Trunk activation and mobility | Deep abdominal activation: 10 × 5 s<br>Supine pelvic tilt: 10–15 reps<br>Cat–camel: 10 reps | At least 10–15 min/session, or longer as tolerated, according to individual well-being. |
| Weeks 3–5 | Strength and movement control | Bridge: 3 × 10<br>Lateral band walk: 3 × 10 steps<br>Quadriceps stretch: 4 × 5–10 s | At least 15–20 min/session, or longer as tolerated, according to individual well-being. |
| Weeks 6–8 | Functional control and maintenance | Modified side plank: 3 × 10–15 s<br>Hip flexor and hamstring stretches: 2 × 30 s/side/stretch<br>Hip hinge: 2–3 × 8–10 reps | At least 15–20 min/session, or longer as tolerated, according to individual well-being. (maintenance) |

### Common guidance for all supervised sessions

Exercises were performed in a controlled and comfortable manner rather than at maximal effort. Participants were instructed to maintain trunk and pelvic control. If an exercise provoked pain, the range of motion or load was reduced to a pain-free or nearly pain-free level. Exercise difficulty was progressed individually while maintaining movement control.

### Home exercise

The exercises introduced during the supervised sessions were continued at home during the intervening weeks. Participants were instructed to complete the home-based exercises at least three times per week.

### Aerobic exercise

Participants were instructed to exercise for a minimum of 10-15 minutes per session during weeks 1-2 and for a minimum of 15-20 minutes per session from week 3 onwards. Exercise was performed at least three times weekly, with no upper limit on the number of weekly sessions.

**Appendix Table 1.** The 8-week health coaching programme.

| Week | Main content | Monitoring and progression |
| --- | --- | --- |
| 1 | Pain education, goal setting and initiation of physical activity | NRS, PSFS, biopsychosocial assessment, activity goals |
| 2 | Fear avoidance and graded exposure | NRS, PSFS, adherence, avoided activities and exposure targets |
| 3 | Self-management and pain-management strategies | NRS, PSFS, activity progression, pacing, body awareness |
| 4 | Independent practice | Continuation of agreed activities and home exercises |
| 5 | Increasing physical activity and functional capacity | NRS, PSFS, daily activity, symptom fluctuations, exercise adherence |
| 6 | Work-related loading and activity management | NRS, PSFS, work-related loading, graded exposure, load management and movement strategies |
| 7 | Maintenance of physical activity and relapse prevention | Physical activity, fear avoidance, barriers and solutions, continuation plan |
| 8 | Independent continuation | Continued implementation of physical activity, exercise and self-management strategies |

## Appendix 2. The 8-week health coaching programme

The 8-week health coaching programme was based on a biopsychosocial approach to chronic low back pain and aimed to support self-management, reduce fear-avoidance behaviour, and improve physical activity and functional capacity (Table 1). Approximately 30-minute telephone coaching sessions were conducted during weeks 1, 2, 3, 5, 6, and 7. Between the coaching sessions, participants continued individually agreed activities and home exercises.

The coaching focused on enhancing participants’ understanding of chronic low back pain, promoting self-management skills, and supporting gradual increases in physical activity. Early sessions emphasized individualized goal setting, identification of personally meaningful activities, and development of realistic action plans. Participants were encouraged to incorporate physical activity into everyday routines through small, achievable behavioural changes and regular home-based exercises.

Subsequent sessions addressed fear-avoidance beliefs, graded exposure to avoided movements and activities, pain-management strategies, pacing, body awareness, and methods for increasing confidence in movement. Participants were guided to monitor their activity levels, recognize barriers to participation, and identify practical solutions to support adherence.

Later sessions focused on increasing physical activity and functional capacity, integrating movement into work and daily-life situations, and developing sustainable activity habits. Physical activity was encouraged through individually tailored home exercises, daily movement, and gradual progression according to participants’ abilities and goals. Particular attention was paid to reducing fear-related movement avoidance through education, reassurance regarding the safety of movement, and confidence-building strategies.

Week 6 focused specifically on work-related and daily-life loading situations, graded exposure, load regulation, and practical strategies to increase movement throughout the day. During week 7, participants reviewed their progress, identified factors supporting long-term adherence, and developed an individualized continuation plan including concrete goals, anticipated barriers, and strategies for independent self-management after the intervention.

A structured coaching form was used to guide the sessions and monitor individual progress. Pain at rest and during movement was monitored using a Numeric Rating Scale (NRS, 0-10), together with patient-specific functional activities (Patient-Specific Functional Scale, PSFS), physical activity, agreed goals, and progression of self-management strategies.

